# Impact of age and SARS-CoV-2 breakthrough infection on humoral immune responses after three doses of COVID-19 mRNA vaccine

**DOI:** 10.1101/2022.08.08.22278494

**Authors:** Francis Mwimanzi, Hope R. Lapointe, Peter K. Cheung, Yurou Sang, Fatima Yaseen, Rebecca Kalikawe, Sneha Datwani, Laura Burns, Landon Young, Victor Leung, Siobhan Ennis, Chanson J. Brumme, Julio S.G. Montaner, Winnie Dong, Natalie Prystajecky, Christopher F. Lowe, Mari L. DeMarco, Daniel T. Holmes, Janet Simons, Masahiro Niikura, Marc G. Romney, Zabrina L. Brumme, Mark A. Brockman

**Author notes:** **Corresponding Author Contact Information:** Mark A. Brockman, Ph.D., Professor, Faculty of Health Sciences, Simon Fraser University, 8888 University Drive, Burnaby, BC, Canada, V5A 1S6. MAB, MGR and ZLB contributed equally.

## Abstract

**Background:** Longer-term immune response data after three doses of COVID-19 mRNA vaccine remain limited, particularly among older adults and following Omicron breakthrough infection.

**Methods:** We quantified wild-type- and Omicron-specific serum IgG levels, ACE2 displacement activities and live virus neutralization up to six months post-third dose in 116 adults aged 24-98 years who remained COVID-19-naïve or experienced their first SARS-CoV-2 infection during this time.

**Results:** Among 78 participants who remained COVID-19-naïve throughout follow-up, wild-type- and Omicron BA.1-specific IgG concentrations were comparable between younger and older adults, though BA.1-specific responses were consistently significantly lower than wild-type-specific responses in both groups. Wild-type- and BA.1-specific IgG concentrations declined at similar rates among COVID-19-naïve younger and older adults, with median half-lives ranging from 69-78 days. Antiviral antibody function declined substantially over time in COVID-19-naïve individuals, particularly older adults: by six months, BA.1-specific neutralization was undetectable in 96% of older adults, versus 56% of younger adults. SARS-CoV-2 infection, experienced by 38 participants, boosted IgG levels and neutralization above those induced by vaccination alone. Nevertheless, BA.1-specific neutralization remained significantly lower than wild-type, with BA.5-specific neutralization lower still.

**Conclusions:** Our findings underscore the immune benefits of third COVID-19 mRNA vaccine doses in adults of all ages, but rapid decline of Omicron-specific neutralization activity in COVID-19-naïve individuals, particularly among older adults, demonstrates the need for fourth doses within 3-6 months to maintain systemic responses. Individuals who experienced SARS-CoV-2 breakthrough infection post-third vaccine dose however can likely delay a fourth dose beyond this timeframe.

## INTRODUCTION

Third COVID-19 mRNA vaccine doses have been provided to immunocompromised and clinically-vulnerable individuals to complete their initial vaccine series [1-3] and to the general population as “booster doses” to offset the natural decline of systemic antibodies [4, 5] and to augment responses against recent Omicron variants [6] that are more immune-evasive [7-14]. While third doses can enhance vaccine-induced protection against severe disease [1, 15, 16], they are not as effective to prevent Omicron-driven infections [17-19]. Few studies have assessed the durability of immune responses after three vaccine doses across the adult age spectrum, nor compared vaccine-induced responses to hybrid responses elicited in the now substantial number of individuals who experienced their first SARS-CoV-2 infection post-third vaccine dose, in one of the recent Omicron-driven waves. Longer-term monitoring of antibody responses and antiviral functions specific to both wild-type and Omicron variants among vaccinated individuals who remain COVID-19-naive versus those who experienced breakthrough Omicron infection can help inform the design, delivery and timing of additional COVID-19 vaccine doses.

In a prior study, we reported that older adults mounted weaker antibody responses than younger adults after two mRNA vaccine doses, but initial responses after three doses were equivalent between these groups [20]. Here, we longitudinally examine wild-type- and Omicron (BA.1, BA.2, BA.3 and BA.5)-specific antibody concentrations and/or antiviral function up to six months post-third dose in 116 individuals ranging in age from 24 to 98 years old who remained COVID-19-naïve until at least one month after receiving their third dose. While two-thirds of the cohort remained COVID-19-naïve throughout follow-up, one-third experienced their first SARS-CoV-2 infection during this time, presumably with Omicron BA.1 or BA.2, based on local molecular epidemiology trends during the study period [21]. This allowed us to additionally compare vaccine-induced and “hybrid” (combined vaccine and infection-induced [22]) immune responses specific to wild-type and Omicron variants, across age groups.

## METHODS

### Participants

Our cohort, based in British Columbia (BC) Canada, has been described previously [20]. Here, we studied the subset of 69 healthcare workers (HCW) and 47 older adults (OA) who remained COVID-19 naive until at least one month after their third COVID-19 mRNA vaccine dose (**Table 1**). SARS-CoV-2 infections were detected by the development of serum antibodies against Nucleocapsid (N) using the Elecsys Anti-SARS-CoV-2 assay (Roche Diagnostics), combined with diagnostic (PCR- and/or rapid-antigen-test-based) information where available.

**Table 1:** Participant characteristics.

| Characteristic | HCW<br>(n=69) | Older Adults<br>(n=47) |
| --- | --- | --- |
| **Sociodemographic and health variables** |  |  |
| Age in years, median [IQR] | 40 [34-51] | 78 [72-83] |
| Female sex at birth, n (%) | 53 (77%) | 34 (72%) |
| white Ethnicity, n (%) | 32 (46%) | 35 (74%) |
| Number of chronic health conditions, median [IQR] | 0 [0-1] | 1 [0-2] |
| **Vaccine details** |  |  |
| Initial regimen |  |  |
| BNT162b2 - BNT162b2 | 67 (97%) | 38 (81%) |
| mRNA-1273 - mRNA-1273 | 1 (1.5%) | 7 (15%) |
| heterologous mRNA | 1 (1.5%) | 2 (4%) |
| Third dose |  |  |
| BNT162b2 | 33 (48%) | 18 (38%) |
| mRNA-1273 | 36 (52%) | 29 (62%) |
| Date range that third doses were received | Oct 2021-Feb 2022 | Oct 2021-Jan 2022 |
| Time between second and third doses in days, median [IQR] | 210 [199-232] | 198 [173-216] |
| **Specimen collection** |  |  |
| 1 month after third dose, n (%) | 68 (99%) | 47 (100%) |
| 3 months after third dose, n (%) | 68 (99%) | 45 (96%) |
| 6 months after third dose, n (%) | 57 (83%) | 44 (94%) |
| **Post-vaccination SARS-CoV-2 infections, n (%)** | 30 (43%) | 8 (17%) |

### Ethics approval

Written informed consent was obtained from all participants or their authorized decision makers. This study was approved by the University of British Columbia/Providence Health Care and Simon Fraser University Research Ethics Boards.

### Assays

We quantified IgG-binding antibodies in serum against the SARS-CoV-2 Spike Receptor Binding Domain (RBD) using the V-plex SARS-CoV-2 (IgG) ELISA kit (Panel 22; Meso Scale Diagnostics), which features WT (Wuhan) and Omicron (BA.1) RBD antigens, on a Meso QuickPlex SQ120 instrument. Serum was diluted 1:10000, with results reported in Arbitrary Units (AU)/mL. We assessed surrogate virus neutralization activity [23] in serum by competition ELISA using the same kit (Panel 22; V-plex SARS-CoV-2 [ACE2]) to measure blockade of the RBD-ACE2 receptor interaction. Sera were diluted 1:40 and results reported as the percentage (%) ACE2 displacement. A subset of specimens was also tested for IgG-binding antibodies and ACE2 displacement using V-plex ELISA kits that featured Wuhan, BA.1, BA.2 and BA.3 Spike antigens (Panel 25; Meso Scale Diagnostics). Virus neutralizing activity in plasma was examined using live wild-type (WT) (USA-WA1/2020; BEI Resources), Omicron BA.1 (GISAID EPI_ISL_9805779) and Omicron BA.5 (GISAID EPI_ISL_15226696) SARS-CoV-2 strains on VeroE6-TMPRSS2 (JCRB-1819) target cells [20]. Virus stocks were diluted to 50 TCID_50_/200 μl in the presence of serial 2-fold plasma dilutions (1/20 to 1/2560) and added to target cells in triplicate. Viral cytopathic effect (CPE) was recorded three days post-infection. Neutralization was reported as the highest reciprocal dilution able to prevent CPE in all three wells. Partial or no neutralization at 1/20 dilution was considered below the limit of quantification (BLOQ).

## RESULTS

### Participant characteristics

HCW and OA were a median of 40 and 78 years old, respectively (overall range 24-98 years old), and predominantly female (**Table 1**). OA were predominantly of white ethnicity (74%, compared to 46% of HCW) and had more chronic health conditions (median of 1, interquartile range [IQR] 0-2 in OA versus 0 [IQR 0-1] in HCW). Most participants (97% of HCW and 81% of OA) initially received two doses of BNT162b2; the remainder received two doses of mRNA-1273 or a heterologous mRNA vaccine regimen. Third doses were predominantly mRNA-1273 (52% of HCW and 62% of OA), which were administered in an age-dependent manner per local guidelines: most OA received 100 mcg whereas HCW received 50 mcg. Third doses were administered an average of ∼7 months after the second dose. During follow-up, 43% of HCW and 17% of OA experienced their first SARS-CoV-2 infection, the vast majority of which were likely Omicron BA.1 or BA.2, based on local molecular epidemiology trends during the study period [21].

### Binding antibody responses after vaccination and breakthrough Omicron infection

As reported previously [20, 24], anti-RBD serum IgG concentrations specific to both WT and Omicron BA.1 were significantly lower in COVID-19-naïve OA compared to younger HCW after two vaccine doses, but the third dose substantially enhanced IgG responses in both groups, such that they reached equivalence at one month post-third-dose (**Figure 1A**). At this time point for example, WT-specific anti-RBD IgG concentrations were a median of 5.21 (IQR 5.08-5.45) log_10_ AU/mL in HCW versus 5.25 (IQR 4.99-5.43) log_10_ AU/mL in OA (p=0.7). Among the participants who remained COVID-19-naive throughout follow-up, WT-specific IgG responses declined to a median 4.95 (IQR 4.79-5.16) log_10_ AU/mL in HCW versus 4.96 (IQR 4.76-5.17) log_10_ AU/mL in OA at three months post-third dose, levels that were not significantly different between groups (p=0.9). By six months post-third dose, WT-specific IgG responses had declined to a median 4.64 (IQR 4.42-4.84) log_10_ AU/mL in HCW versus 4.59 (IQR 4.35-4.87) log_10_ AU/mL, levels that were again not significantly different between groups (p=0.7). Nevertheless, the magnitude of these declines meant that WT-specific IgG concentrations in COVID-19-naïve HCW had declined to significantly below peak post-second-dose levels by six months after the third dose, (p<0.0001), while in OA they had declined to comparable levels as those elicited by two doses (p=0.2). Identical trends were observed for BA.1-specific IgG among COVID-19-naïve HCW and OA, though these were on average ∼0.6 log_10_ AU/mL lower than WT-specific responses at all time points, differences that were statistically significant at p<0.0001 in all cases (though these comparisons are not shown in Figure 1A to minimize clutter).

**Figure 1.**
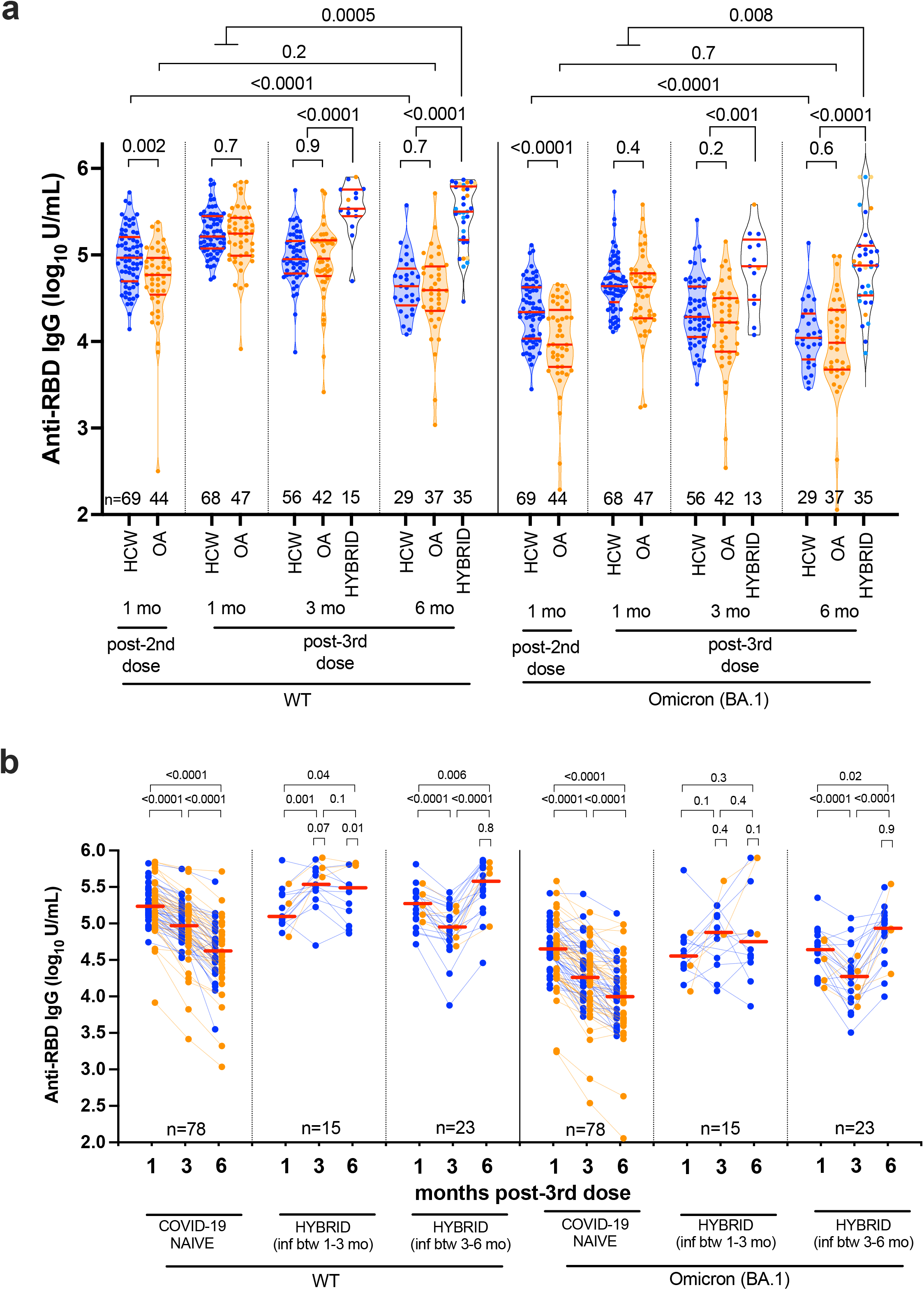
WT- and Omicron BA.1-specific anti-RBD IgG concentrations following three-dose COVID-19 vaccination. *Panel A:* Longitudinal serum anti-RBD IgG concentrations specific to WT (left side) and Omicron BA.1 (right side) in COVID-19-naive health care workers (HCW; blue circles) and older adults (OA; orange circles). Any participant who experienced a SARS-CoV-2 breakthrough infection between 1-3 or 3-6 months post-third-dose was reclassified into the “hybrid” group at their following study visit, with HCW in blue and OA in orange circles. At six months post-third-dose, the darker-colored symbols in the hybrid group denote recent infections that occurred between 3-6 months, while the lighter-colored symbols denote the infections that had previously occurred between 1-3 months. Red bars indicate median and IQR. Comparisons between independent groups were performed using the Mann-Whitney U-test; longitudinal paired comparisons were performed using the Wilcoxon matched pairs test. P-values are not corrected for multiple comparisons. *Panel B:* Same data as panel A, but where serum anti-RBD IgG concentrations are plotted longitudinally by participant (HCW in blue, OA in orange). Participants are stratified into three groups: those who remained COVID-19 naive throughout the study, those who acquired SARS-CoV-2 between 1-3 months post-third-dose, those who acquired SARS-CoV-2 between 3-6 months post-third-dose. Horizontal red lines denote the overall median response at each time point, where HCW and OA are treated as a combined group. P-values on top of larger brackets compare responses between time points using the Wilcoxon matched pairs test, where HCW and OA are treated as a combined group. P-values on top of small brackets compare responses between HCW and OA at each time-point post SARS-CoV-2 infection using the Mann-Whitney U test. P-values are not corrected for multiple comparisons.

By contrast, individuals who experienced their first SARS-CoV-2 infection at some time between one and six months following their third vaccine dose exhibited markedly higher virus-specific IgG concentrations than their COVID-19-naïve counterparts at both three and six months post-third dose (all comparisons p<0.001 for both WT- and BA.1-specific responses; **Figure 1A**). Moreover, stratifying the hybrid immune responses by participant age group revealed that older adults who contracted SARS-CoV-2 mounted equivalent or higher WT-specific IgG responses compared to younger HCW at all time points studied (p-values ranging from 0.01 to 0.8), and equivalent BA.1-specific IgG responses compared to HCW (p-values ranging from 0.1-0.9) (**Figure 1B**). In fact, at six months post-third-dose, both WT- and BA.1-specific IgG concentrations among the hybrid immune participants of all ages were significantly higher than their original peak responses induced by three vaccine doses alone: at six months the median WT-specific IgG concentration was 5.50 (IQR 5.17-5.79) log_10_ AU/mL in the combined hybrid group, which was ∼0.25 log_10_ AU/mL higher than that observed at one month post-third dose (p=0.0005). Visualizing the data in terms of individual-level longitudinal responses revealed that, while most individuals enjoyed a substantial boost in terms of both WT- and BA.1-specific responses following SARS-CoV-2 infection, responses in a minority of participants remained constant or even declined thereafter (**Figure 1B)**.

Our longitudinal data also allowed us to estimate the half-lives of wild-type and BA.1-specific IgG concentrations in serum among individuals who remained COVID-19 naive over the subsequent six months. Following three vaccine doses, the half-life of WT-specific IgG was estimated to be a median 73 (IQR 53-101) days in COVID-19-naïve HCW versus 69 (IQR 54-91) days in OA, a difference that was not statistically significant (p=0.8; **Figure S1A, B**). In COVID-19-naïve individuals, BA.1-specific IgG half-lives were similar to those for WT and were also comparable between OA (median 75 days [IQR 58-93]) and HCW (78 [IQR 64-94]) (p=0.5; **Figure S1C, D**).

### ACE2 displacement activity after vaccination and breakthrough Omicron infection

As reported previously [20, 24], the antiviral function of antibodies measured by ACE2 displacement activity against WT and the Omicron BA.1 variant was lower in COVID-19-naïve OA compared to HCW following two vaccine doses, but these responses increased to equivalence one month after the third dose. Specifically, these responses reached a median 98.7% (IQR 96.3-99.3) in HCW versus 99.3% (IQR 96.0-99.7) OA for WT (p=0.12), and a median of 62.5% (IQR 46.2-75.5) in HCW versus 66.2% (IQR 44.6-79.3) in OA for BA.1 (p=0.4) (**Figure 2A**). Among the participants who remained COVID-19-naive throughout follow-up, WT-specific ACE2 displacement activities declined similarly in both groups, to a median 98.0% (IQR 93.7-99.3) in HCW versus 98.3% (IQR 91.7-99.5) in OA at three months after the third dose (p=0.9) and a median 92.8% (IQR 80.0-99.9) in HCW versus 91.4% (IQR 72.3-97.1) in OA at six months (p=0.4). Somewhat in contrast, BA.1-specific ACE2 displacement activity declined slightly faster in COVID-19-naïve OA compared to HCW, particularly between three and six months after the third dose. Specifically, BA.1-specific ACE2 displacement activities had declined to a median 48.6% (IQR 20.5-73.8) in HCW versus 47.6% (IQR 14.2-71.7) in OA at three months (p=0.4) and to a median 43.3% (IQR 13.4-62.2) in HCW versus 20.7% (IQR 5.6-32.3) in OA at six months (p=0.04). Similar to the declines observed in IgG concentrations over time, by six months post-third dose, WT- and BA.1-specific ACE2 displacement activities in COVID-19-naïve HCW had declined to below the peak levels elicited after only two vaccine doses (p<0.0001 and p=0.02 respectively), while in COVID-19-naïve OA they had declined to comparable levels as achieved after two doses (p=0.6 and p=0.4, respectively).

**Figure 2.**
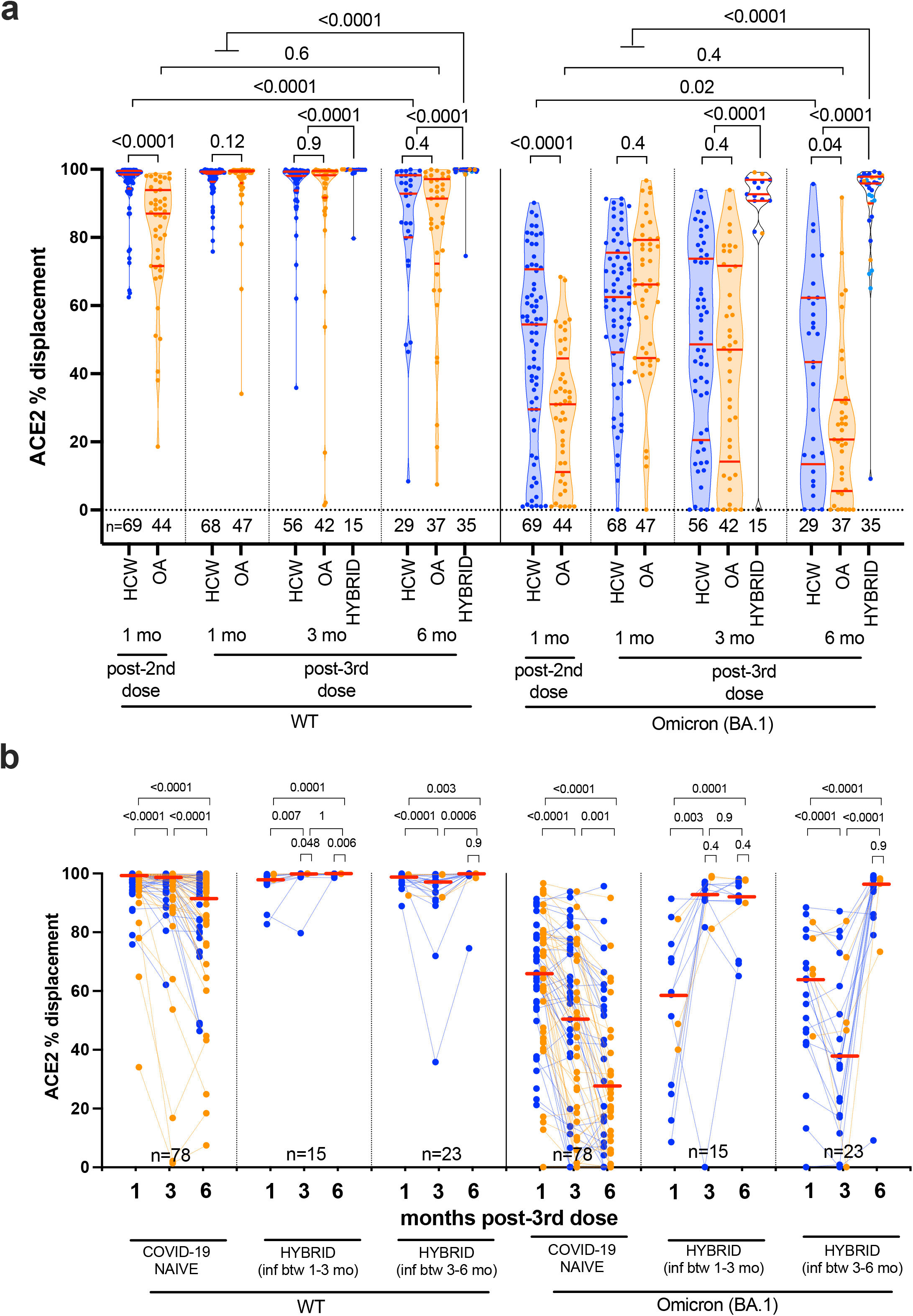
WT- and Omicron BA.1-specific ACE2% displacement function following three-dose COVID-19 vaccination. *Panel A:* Same as Figure 1A, but for ACE2 displacement activity in serum, a surrogate measure of virus neutralization, where results are reported in terms of % ACE2 displacement. *Panel B:* Same data as panel A, but where ACE2 % displacement activities are plotted longitudinally by participant (HCW in blue, OA in orange). The legend is the same as for Figure 1B.

By contrast, individuals who experienced their first SARS-CoV-2 infection between one and six months following their third vaccine dose showed a strong boost in ACE2 displacement activity at both three and six months, where this difference was most apparent for the BA.1-specific responses (all comparisons p<0.0001 for both WT and BA.1; **Figure 2A**). In fact, at six months after the third dose, both WT- and BA.1-specific ACE2 displacement activities in the hybrid immune group significantly exceeded those induced by vaccination alone (both comparisons p<0.0001; **Figure 2A**). For example, BA.1-specific ACE2 displacement activities rose to a median 95.9% (IQR 90.0-97.8) in the hybrid group at six months, levels that were an average 30% higher than those induced by three-dose vaccination alone. Similar to the results observed for IgG concentrations, stratifying the hybrid immune responses by participant age group revealed that older adults who contracted SARS-CoV-2 exhibited equivalent or higher WT-specific ACE2 displacement activities compared to younger HCW at all time points studied (p-values ranging from 0.006 to 0.9), and equivalent BA.1-specific ACE2 displacement activities compared to HCW (p-values ranging from 0.4-0.9) (**Figure 2B**). Also similar to the IgG observations, visualizing the ACE2 displacement results in terms of individual-level longitudinal responses revealed that SARS-CoV-2 infection enhanced both WT- and Omicron BA.1-specific ACE2 displacement activities in most, but not all, individuals (**Figure 2B)**.

### Virus neutralization activity after vaccination and breakthrough Omicron infection

As reported previously [20, 24], live virus neutralization activities against both the WT and Omicron BA.1 variants were weaker in OA compared to younger HCW following two vaccine doses, but these activities were enhanced and reached equivalence one month after the third dose (**Figure 3A**). Specifically, at this time point, the reciprocal plasma dilutions required to neutralize WT were a median 320 (IQR 160-320) in HCW versus a median 320 (IQR 80-640) in OA (p=0.9), while those required to neutralize BA.1 were a median 40 (IQR 20-80) in both groups (p=0.8). Among the participants who remained COVID-19-naïve throughout follow-up, WT neutralization activity declined relatively similarly in both groups, to median reciprocal dilutions of 80 (IQR 80-160) in HCW versus 80 (IQR 40-160) in OA at three months post-third dose (p=0.03), and a median 40 (IQR 20-80) in HCW versus 20 (IQR 20-80) in OA at six months (p=0.3). Nevertheless, by six months, WT-specific neutralization activities in COVID-19-naïve HCW and OA had declined to below the peak levels induced by two vaccine doses (p<0.0001 in HCW and p=0.045 in OA; **Figure 3A**). Moreover, BA.1-specific neutralization activity declined faster in COVID-19-naïve OA compared to HCW after three vaccine doses. Specifically, at three months post-third-dose, BA.1-specific neutralization in HCW had declined to a median reciprocal dilution of 20 (IQR 20-40), whereas in 79% of OA this function was below the limit of quantification (BLOQ) (p=0.004). By six months, BA.1-specific-neutralization activity had declined to BLOQ in 56% and 96% of COVID-19-naive HCW and OA respectively (p=0.003), activities that were lower than peak responses induced by two vaccine doses (p=0.02 for HCW; p=0.06 for OA).

**Figure 3.**
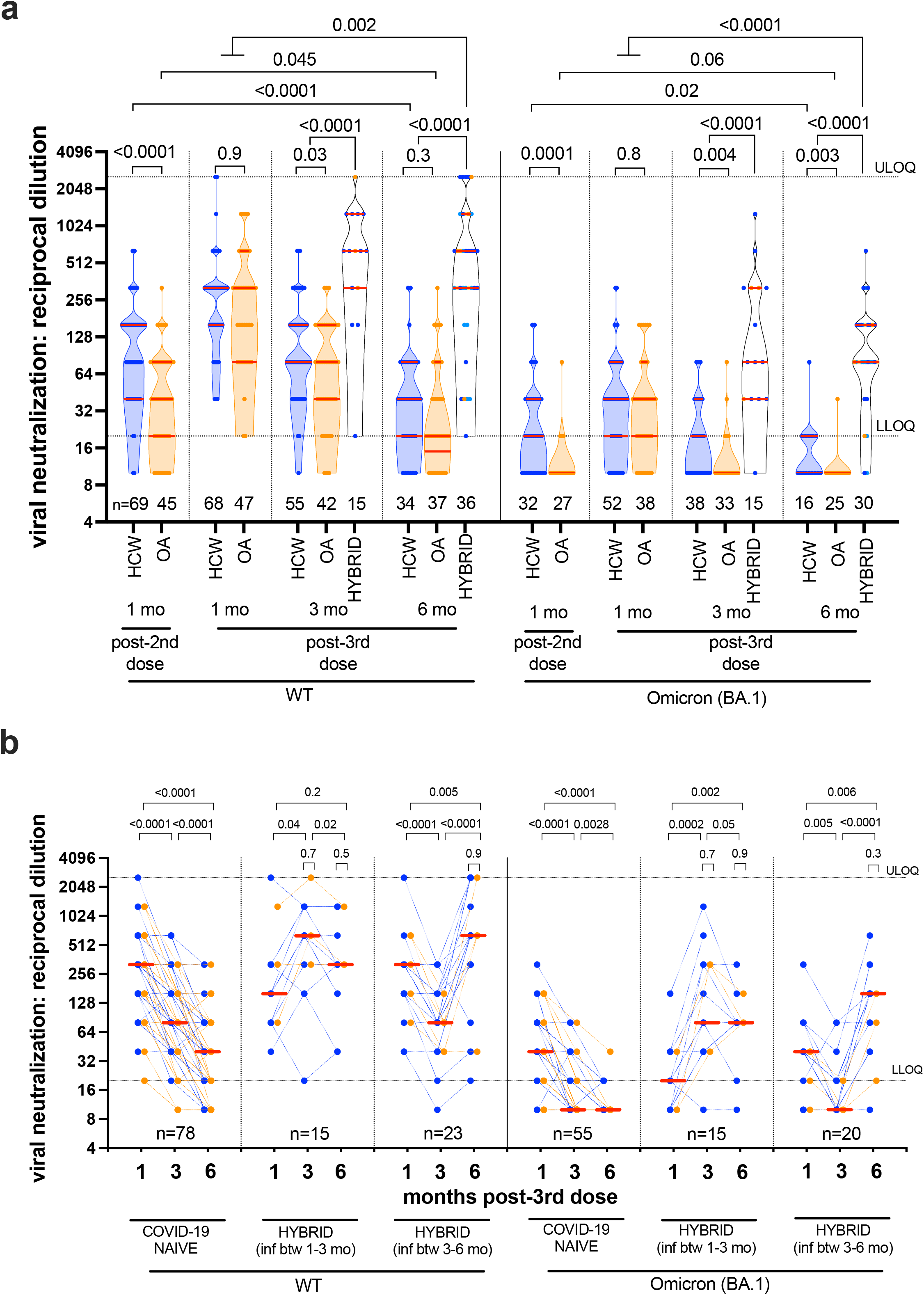
WT- and Omicron BA.1-specific live virus neutralization activity following three-dose COVID-19 vaccination. *Panel A:* Same as Figure 1A, but for live virus neutralization activity, defined as the lowest reciprocal plasma dilution at which neutralization was observed in all wells of a triplicate assay. Serial two-fold dilutions of 1/20 (lower limit of quantification; LLOQ) to 1/2560 (upper limit of quantification; ULOQ) were tested. Plasma samples showing neutralization in fewer than three wells at a 1/20 dilution are displayed as a reciprocal dilution of “10” and were reported as below limit of quantification (BLOQ) in the text. Omicron-BA.1 specific neutralization was performed on a subset of samples only. *Panel B:* Same data as panel A, but where neutralization activities are plotted longitudinally by participant (HCW in blue, OA in orange). The legend is the same as for Figure 1B. Note that many datapoints are superimposed in both panels.

By contrast, individuals who experienced their first SARS-CoV-2 infection between one and six months following their third vaccine dose showed a strong enhancement in their ability to neutralize both WT and BA.1 (all comparisons p<0.0001; **Figure 3A**). At six months, participants in the hybrid immune group displayed significantly higher WT- and BA.1-specific neutralization activities compared to peak responses induced by three vaccine doses alone (all p≤0.002; **Figure 3A**): at this time point BA.1-specific neutralization in the hybrid immune group was achieved with a median reciprocal dilution of 80 (IQR 80-160), compared to a median 40 (IQR 20-80) observed in the combined HCW and OA group at one month following the third vaccine dose.

Similar to observations made for other humoral responses, stratifying the hybrid immune responses by age indicated that OA exhibited equivalent WT- and BA.1-specific neutralization activities compared to younger HCW at all time points (p-values ranging from 0.3 to 0.9) (**Figure 4B**). Furthermore, visualization of individual-level longitudinal responses revealed that while SARS-CoV-2 infection boosted both WT- and BA.1-specific neutralization activities in most individuals, some individuals’ responses remained stable or declined following infection (**Figure 4B**).

**Figure 4.**
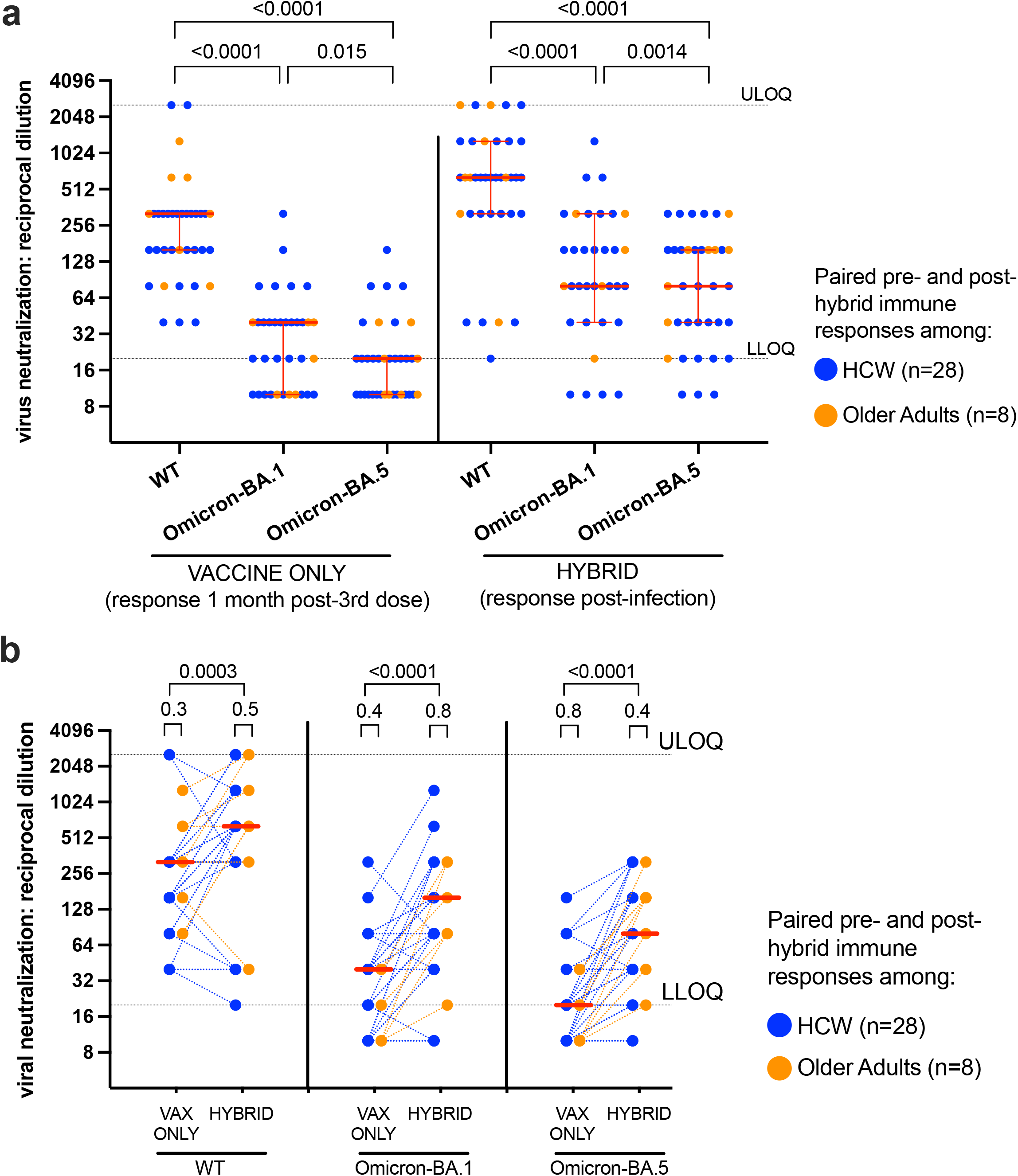
WT-, Omicron BA.1- and Omicron BA.5-specific live virus neutralization activity before and after attainment of hybrid immunity. *Panel A:* WT-, Omicron BA.1- and Omicron BA.5-specific neutralization activities in 28 HCW (blue circles) and 8 OA (orange circles) one month after the 3rd vaccine dose (“vaccine only”) and after subsequent SARS-CoV-2 infection (“hybrid”). In this panel, data are plotted to facilitate comparisons between neutralization activities to the three SARS-CoV-2 variants, before and after attainment of hybrid immunity. P-values, computed on combined HCW and OA as a combined group, were computed using the Wilcoxon matched pairs test. P-values are not corrected for multiple comparisons. ULOQ/LLOQ: upper/lower limit of quantification. *Panel B*: Same data as in panel A, but plotted longitudinally by participant, and stratified by SARS-CoV-2 variant, to highlight the change in neutralization activity before and after attainment of hybrid immunity. Red lines indicate medians of HCW and OA treated as a combined group. P-values on top of large brackets compare responses before and after hybrid immunity using the Wilcoxon matched pairs test. P-values on top of small brackets compare responses between HCW and OA at a given time point using the Mann-Whitney U test. Note that a large number of points are superimposed in this plot.

### Responses to newer Omicron variants, including BA.5

While the BA.1 variant drove the first global wave of Omicron infections, this strain has largely been replaced by newer Omicron variants that demonstrate distinct abilities to evade neutralizing antibodies [8, 25-29]. To begin to evaluate vaccine and hybrid immune responses to other Omicron strains, we quantified anti-Spike IgG and ACE2 displacement activities specific to WT, BA.1, BA.2 and BA.3 variants in a subset of 25 COVID-19 naïve participants at one month after their third vaccine dose (**Figure S2)**. Compared to WT-specific responses, anti-Spike IgG concentrations were on average ∼0.4 log_10_ AU/mL lower for each of these Omicron variants, while ACE2 displacement activities were on average at least 30% lower for these Omicron variants (all p<0.0001; **Figure S2**). BA.2-specific anti-Spike IgG concentrations were comparable to BA.1 (p=0.2), while BA.2-specific ACE2 displacement activities were marginally lower (p=0.01). BA.3-specific anti-Spike IgG concentrations and ACE2 displacement activities were significantly lower than those against BA.1 (both p<0.0001), though the magnitude of these differences was modest (*e*.*g*. median IgG was 5.42 log_10_ AU/mL for BA.1 versus 5.35 log_10_ AU/mL for BA.3). The correlation between the present anti-Spike and the original anti-RBD measurements for the WT and BA.1 antigens (shown in Figure 1) was strong (Spearman’s ρ≥0.86, p<0.0001; **Figure S2**).

Omicron BA.5 emerged in early 2022 [30] and is currently the dominant SARS-CoV-2 variant circulating globally [31-34]. To investigate vaccine- and hybrid-induced immune responses against this strain, we performed live virus neutralization assays using a local BA.5 isolate in a subset of 36 participants (28 HCW and 8 OA) who experienced SARS-CoV-2 breakthrough infection between one and six months after receiving three vaccine doses (**Figure 4)**. For each individual, vaccine-induced neutralization activity was tested at one-month post-third dose, and hybrid-induced neutralization activity was tested following SARS-CoV-2 anti-N seroconversion (either three months or six months post-third vaccine dose). Based on local molecular epidemiology reports [21], SARS-CoV-2 infections among these participants were likely due to BA.1 or BA.2 (not BA.5). Neutralization results for BA.5 were compared to those against WT and BA.1 at the same timepoints (*i*.*e*. the data from Figure 4). Consistent with recent reports showing that BA.5 is more immune-evasive than prior Omicron variants [26-28], we observed that neutralization activity against BA.5 was significantly lower than activities against WT and BA.1 following three vaccine doses (p<0.0001 and p=0.015, respectively) (**Figure 4A**). Indeed, the median reciprocal dilution for BA.5 neutralization was 20 (IQR BLOQ-20) at this time point. Neutralization activity against all three virus strains was enhanced significantly following infection (all p≤0.0003) (**Figure 4B**), but the hybrid immune response against BA.5 (median reciprocal dilution 80, IQR 40-160) nevertheless remained significantly lower than that against both WT (p<0.0001) and BA.1 (p=0.0014) (**Figure 4A**). Overall, no significant difference in BA.5 neutralization activity was found between OA and younger HCW at either one month post-third vaccine dose nor after acquiring hybrid immunity (**Figure 4B**).

## DISCUSSION

Our observations confirm that a third COVID-19 mRNA vaccine dose significantly enhances antibody responses against both WT and Omicron variants in COVID-19-naïve individuals, particularly among older adults. Our results also shed light on the durability of these responses, revealing that WT- and Omicron BA.1-specific anti-RBD binding IgG concentrations remained comparable in magnitude and declined at similar rates in older and younger adults in the six months following the third dose, though BA.1-specific responses were consistently ∼0.6 log_10_ AU/mL lower than those against WT. By contrast, antiviral antibody functions, particularly those specific to Omicron, declined substantially in all participants who remained COVID-19-naïve over the study period, and especially so in older adults. By six months post-third-dose, antibody responses in COVID-19-naïve participants of all ages had declined markedly to or below the peak levels elicited by two vaccine doses, and BA.1-specific neutralization activity had declined to below the limit of quantification in 56% of younger adults and 96% of older adults. In fact, BA.1-specific neutralization activity had already declined to below the limit of quantification in 79% of COVID-19-naïve older adults by three months after the third dose. These observations, along with the finding that ACE2 displacement function declined more rapidly in COVID-19 naive older adults, suggest that Omicron-specific antibody function may be impaired in older age, but where this impairment is only revealed as antibody concentrations decline.

By contrast, both younger and older adults who experienced their first SARS-CoV-2 infection (presumably Omicron BA.1 or BA.2 [21]) after receiving three vaccine doses demonstrated superior binding antibody concentrations and functional responses, including against the heterologous Omicron variant BA.5, compared to those induced by three vaccine doses alone, though the ability to neutralize BA.5 remained significantly poorer than the ability to neutralize BA.1 even after infection. Nevertheless, and importantly, the magnitude of humoral responses following acquisition of hybrid immunity did not differ significantly between older and younger adults (though the modest number of older adults with hybrid immunity should be acknowledged). These results are consistent with other studies of hybrid immunity [22, 35, 36] and suggest that a post-third-dose Omicron infection will prolong immune protection against Omicron strains for at least a short period. The observation that viral infection led to a pronounced enhancement of Omicron-specific responses, including binding antibody concentration and virus neutralization activity, is likely attributable to exposure to Omicron Spike. If so, bivalent vaccines that include variant Omicron Spike antigens may offer similar advantages, including the ability to elicit superior immune responses against circulating variants compared to existing WT-only vaccines. Intriguingly, while most participants displayed a significant boost in humoral responses following virus infection (and will potentially benefit from hybrid immunity), this did not occur in all cases. Additional research will be necessary to investigate the factors that contribute to such divergent outcomes.

## CONCLUSSIONS

In conclusion, this study underscores the immune benefits of third/booster COVID-19 vaccine doses in all ages. The results also suggest that younger adults who have received three COVID-19 vaccine doses but who have not yet experienced a SARS-CoV-2 infection will require an additional vaccine dose within six months to maintain systemic antibody responses, while SARS-CoV-2-naïve older adults may benefit from an additional vaccine dose within as few as three months. By contrast, individuals who experienced SARS-CoV-2 breakthrough infection after receiving three vaccine doses can likely delay a fourth dose beyond this time frame. Additional studies are needed to assess the durability of hybrid immune responses and to evaluate cross-reactivity against emerging SARS-CoV-2 variants, particularly in the context of bivalent vaccines that incorporate Omicron Spike antigen.

## Data Availability

All data produced in the present study are available upon reasonable request to the authors. Upon study conclusion, data will also be deposited into a national database managed by the funder (CITF). Viral sequences are deposited into GISAID

## AUTHOR CONTRIBUTIONS

MGR, ZLB and MAB led the study. HRL coordinated the study. FM, PKC, YS, FY, LB, SE and WD collected data under the supervision of MLD, VL, JSGM, DTH, JS, MGR, ZLB and MAB. FM, HL, MAB and ZLB analyzed data. YS curated the specimen repository. LY performed phlebotomy and assisted with logistics. FM, YS, RK and SD processed specimens and curated data. CJB advised on data analysis. NP provided data. CFL provided specimens. MN isolated SARS-CoV-2. ZLB and MAB wrote the manuscript. All authors contributed to manuscript review and editing.

## ACKNOWLEDGEMENTS

We thank the leadership and staff of Providence Health Care, including long-term care and assisted living residences, for their support of this study. We thank the phlebotomists and laboratory staff at St. Paul’s Hospital, the BC Centre for Excellence in HIV/AIDS, the Hope to Health Research and Innovation Centre, and Simon Fraser University for assistance. Above all, we thank the participants, without whom this study would not have been possible.

## Supplemental Figures

**Figure S1:**
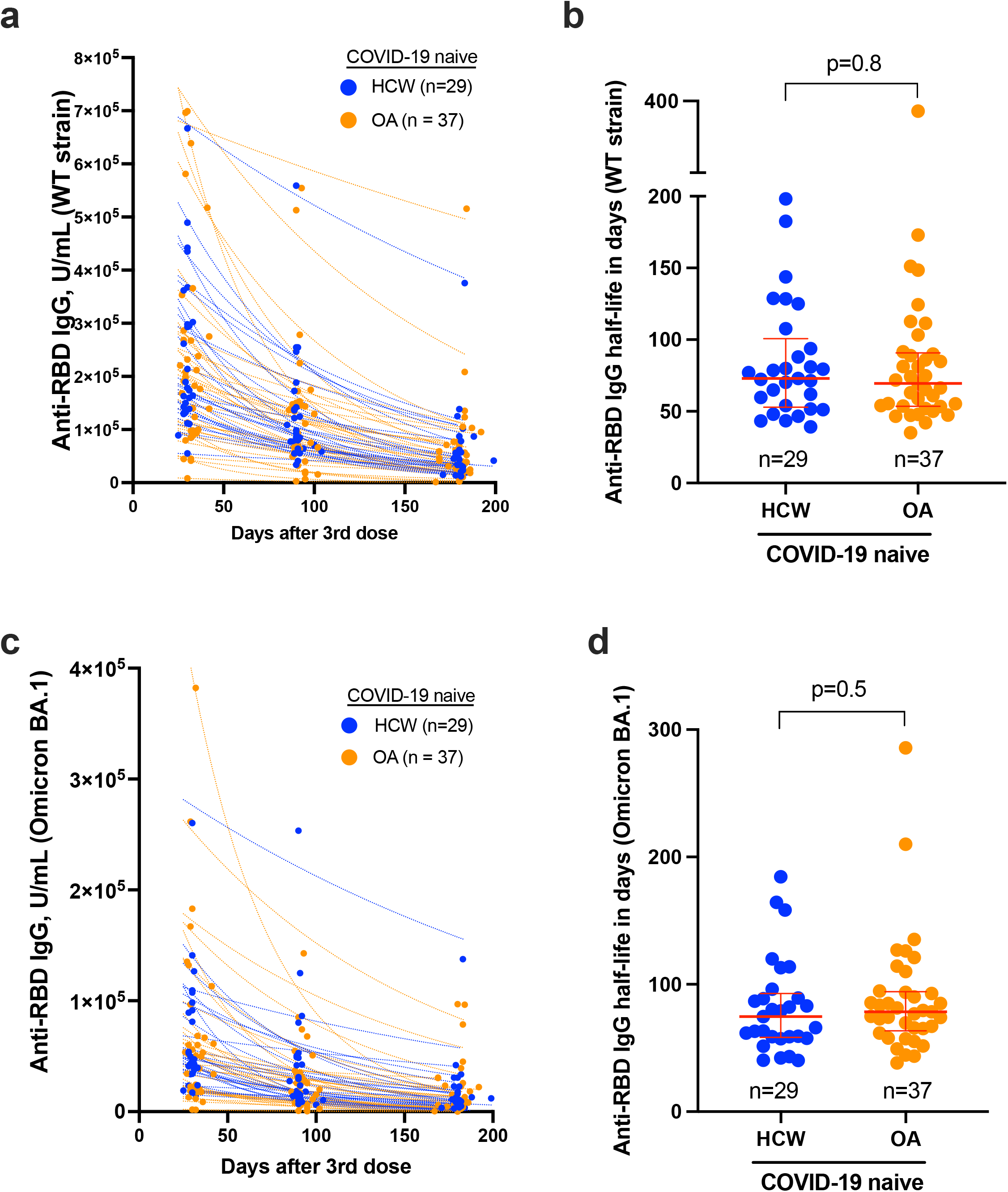
Temporal declines in WT- and Omicron BA.1-specific anti-RBD IgG decline following three-dose COVID-19 vaccination. *Panel A:* Temporal declines in WT-specific anti-RBD IgG responses following three-dose vaccination in HCW (blue) and OA (orange) who remained COVID-19-naive during follow-up, and who completed all three post-third dose visits. *Panel B*: Estimated WT-specific IgG half-lives following three-dose vaccination, calculated by fitting an exponential curve to each participant’s data in panel B. Red bars and whiskers represent the median and IQR. P-values were computed using the Mann-Whitney U-test. *Panel C*: Same as panel A, but for Omicron (BA.1)-specific anti-RBD IgG responses. *Panel D*: Same as panel B, but calculated from the Omicron (BA.1)-specific anti-RBD IgG response data.

**Figure S2:**
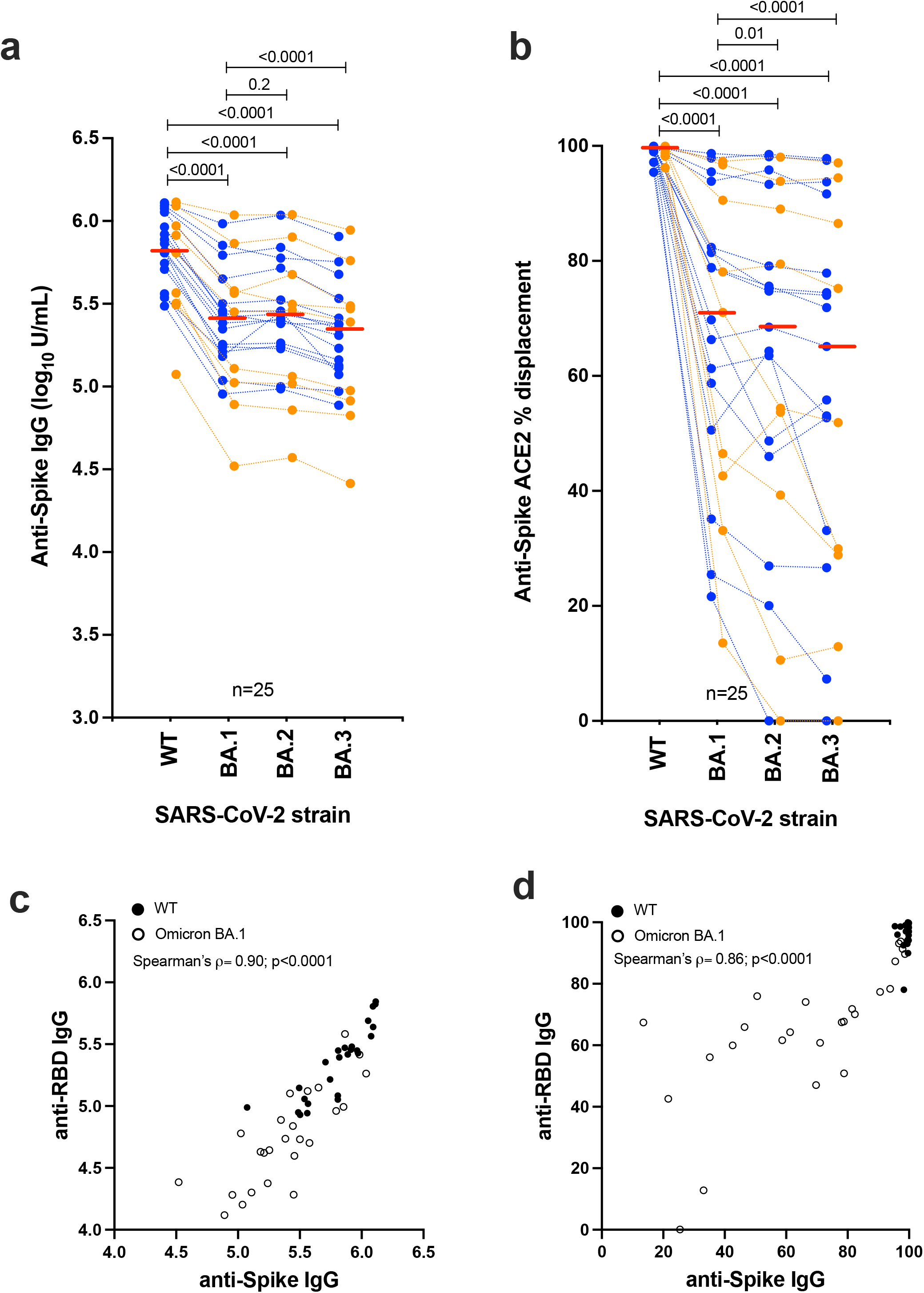
Anti-S IgG and ACE2 displacement activities against additional Omicron variants. *Panel A:* Concentrations of IgG capable of binding WT, Omicron BA.1, BA.2 and BA.3 Spike proteins in a subset of n=25 participants measured one month post-third vaccine dose. HCW in blue; OA in orange; dots are jittered for visualization. P-values are computed using the Wilcoxon matched pairs test and are uncorrected for multiple comparisons. Horizontal red lines denote the overall median response to each variant (HCW and OA are treated as a combined group). *Panel B*: same as Panel A, but for ACE2% displacement activity. *Panel C*: Correlation between WT and Omicron BA.1-specific anti-Spike IgG concentrations (shown in this figure) with the original anti-RBD IgG measurements shown in the primary analysis (in Figure 1A). *Panel D*: Correlation between WT and Omicron BA.1-specific anti-Spike ACE2 displacement with the original anti-RBD measurements shown in the primary analysis.

